# Risk of SARS-CoV-2 infection from contaminated water systems

**DOI:** 10.1101/2020.06.17.20133504

**Authors:** Jamie Shutler, Krzysztof Zaraska, Tom Holding, Monika Machnik, Kiranmai Uppuluri, Ian Ashton, Łukasz Migdał, Ravinder Dahiya

## Abstract

Following the outbreak of severe acute respiratory syndrome coronavirus (SARS-CoV-2) in China, airborne water droplets (aerosols) have been identified as the main transmission route, although other transmission routes are likely to exist. We quantify SARS-CoV-2 virus survivability within water and the risk of infection posed by faecal contaminated water within 39 countries. We identify that the virus can remain stable within water for up to 25 days, and country specific relative risk of infection posed by faecal contaminated water is related to the environment. Faecal contaminated rivers, waterways and water systems within countries with high infection rates can provide infectious doses >100 copies within 100 ml of water. The implications for freshwater systems, the coastal marine environment and virus resurgence are discussed.

## Introduction

The outbreak of the severe acute respiratory syndrome coronavirus (SARS-CoV-2) began in Wuhan province, China in December 2019 and has now spread throughout the world with about 6 million cases confirmed globally within 214 countries and territories. Water aerosols originating from individuals infected by SARS-CoV-2 are considered a major pathway for infection ^1^, and the virus has been shown to remain stable in saline solution ^2^ and under varying environmental conditions ^3^. Viral shedding in faeces of viable SARS-CoV-2 virus is documented (eg ^4^) and SARS-CoV-2 ribonucleic acid (RNA) has been detected in the shed faeces of both symptomatic and asymptomatic children and adults (eg ^5^); with potentially 43% of infections being asymptomatic and unreported ^6^.

Human viral pathogens that can be transmitted by water that pose moderate to high health significance as defined by the WHO include adenovirus, astrovirus, hepatitis A and E, rotavirus, norovirus and other enteroviruses. The survival of the large family of coronavirus in water systems has been highlighted ^7^, and viral loads within untreated wastewater, consistent with population infection rates, have been identified ^8^. While evidence for SARS CoV-2 is limited, other human coronaviruses are documented to survive in wastewater effluent ^9^, with colder water temperature likely to increase survival considerably ^3^. Collectively this evidence suggests that SARS-CoV-2 virus can survive within water and the viral loads within untreated sewage effluent are likely high in countries of high infection rates, a portion of which is viable virus, and therefore water contaminated with sewage provides a potential faecal-oral transmission route (eg ^10^).

Sewage can directly enter natural water systems due to combined sewer overflow events and sewage exfiltration from pipes (eg ^11^) unexpected failure of water treatment systems or a complete lack of water treatment infrastructure, providing a pathway for onward transmission. For example, during the current pandemic large sewage spills, flooding dwellings and community spaces, have occurred in America (within Georgia, Florida and New York) and Spain (Andalucia), while temporary settlements (eg shanty towns, favelas or bustees) and refugee camps are less likely to have safe sanitation systems. Within these settings, this water system pathway could enable viral infection to humans or other susceptible animals via water ingestion or through filtering of water during feeding.

The highly skewed distribution of infected patient viral loads observed ^12^ contain the effects of super spreaders, where single individuals can be responsible for the majority of the viral loading. This viral distribution means that sewage originating from populations that contain super spreaders will contain very high viral loads, even though the majority of the population contribute relatively low viral loadings.

Considering the above, we identify the survivability of SARS-CoV-2 within water systems using published in vitro study data ^3^. We then used an established ‘down the drain’ pollution analysis to calculate the dilution in rivers ^13^, combined with our empirical virus survivability model, to calculate of the relative risk posed to humans by sewage spills within 39 countries. Results using infection numbers on May 03 2020 for 21 countries, where inland water temperatures were available, identify viable waterborne virus concentrations that, if faecal contamination had occurred, would result in a high probability of infection. The implications of these findings for waterborne virus transmission to humans and animals are discussed and recommendations for reducing risk of infection are given.

## Results

Exponential temperature driven survivability identifies that the virus can remain stable and above detection limits for up to 25 days (figure 1a). The relative risk, the normalized country comparable risk associated with a sewage spill after dilution within rivers (figure 1b, 1c) is dependent upon domestic water usage and riverine dilution, where dilution is dependent upon geographical location, relief and weather. Countries with lowest relative risk are those with both high domestic water usage and high dilution (eg Canada, Norway and Venezuela). Highest relative risk results from a combination of low to medium domestic water usage and low dilution (eg Morocco, Spain, Germany). Translating these results to the proportion of the population infected within 21 countries on May 03 2020 identifies the estimated upper and lower limit of viable waterborne virus concentration within the first 24 hours, assuming that a spill occurred (figure 2; uncertainty on the viable virus concentration is ±68% copies L^-1^). Absolute concentrations are higher and will exist for longer within countries with a combination of higher relative risk, colder water and high population infection rates. Assuming infection requires a dose of 100 copies, then a person within the 3 countries with the highest concentrations (Spain, UK, Morocco) who within 24 hours of a spill ingests 100 ml of the contaminated water could receive a total dose >468 copies resulting in a high probability of infection (table 1; full dosage range across all cases is 46 to 3080 copies). 100 ml is the equivalent of 1 to 2 mouthfuls and swimmers can swallow up to 280 ml in a 45 minute swim ^14^. The combination of figure 1a and figure 2a can be used to understand the viable virus concentration after the first 24 hours. The water temperature-controlled virus survivability means that concentrations reduce quickly in Morocco within 24 hours of a spill, whereas the concentrations remain for longer in Spain and the UK where water temperatures are lower (table 1; Figure 2a).

**Table 1.** Viable virus concentration results for the 3 countries for the 3 May 2020 assuming a spill occurred. Median dilution (DF) along with middle^*^ and high^$^ viable to unviable viral ratio (I) results are given to provide a reasonable range of the concentrations within the first 24 hours. ^&^Low DF and high I results enable the extreme range of concentrations to be estimated. Viral survival rates after 24 and 48 hours show how the viable viral concentrations reduce due to temperature driven die off.

| Country | Code | <sup>*</sup> I=1%,<br>median DF,<br>copies L<sup>-1</sup> | <sup>§</sup> I=10%,<br>median DF,<br>copies L<sup>-1</sup> | <sup>&</sup> I=10%,<br>low DF,<br>copies L<sup>-1</sup> | 100 ml dose for<br>case <sup>§</sup> and total<br>range, copies | 24 hour<br>survival<br>, % | 48 hour<br>survival<br>, % |
| --- | --- | --- | --- | --- | --- | --- | --- |
| Spain | SPA | 632 | 6325 | 6325 | 633 (63<sup>*</sup> to 633<sup>&</sup>) | 67 | 45 |
| UK | GBR | 468 | 4682 | 30792 | 468 (47<sup>*</sup> to 3080<sup>&</sup>) | 72 | 52 |
| Morocco | MAR | 459 | 4595 | 25255 | 459 (46<sup>*</sup> to 2526<sup>&</sup>) | 38 | 15 |

**Figure 1.**
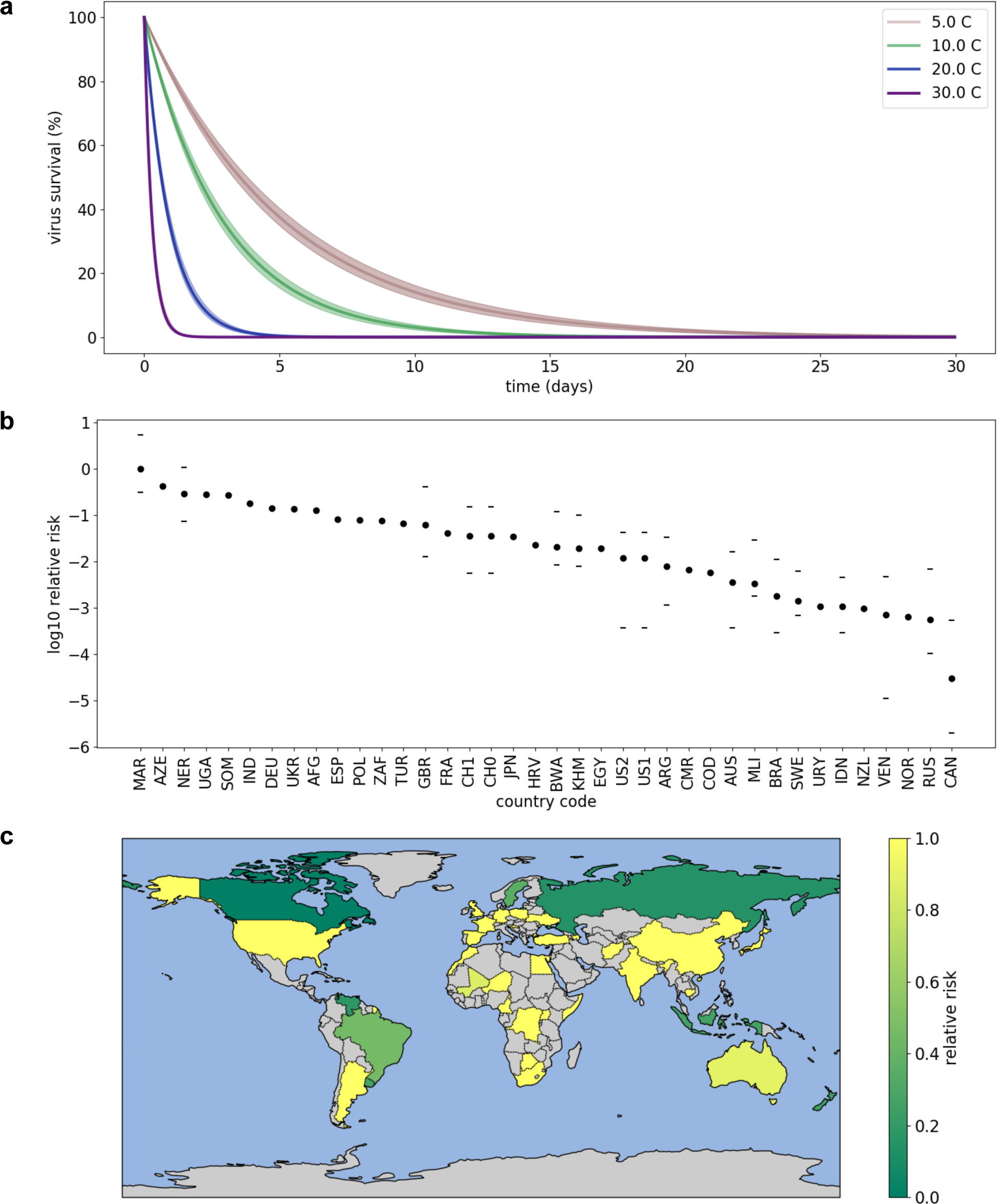
Virus survivability within water and relative risk posed by sewage spills into rivers for 39 countries; a) modelled temperature survivability. Shaded areas show the temperature dependent uncertainties; b) log_10_ relative risk covering the range of 0.001 to 1.0; circles are median values, horizontal lines are 25^th^ and 75^th^ percentiles due to dilution factors from ^13^ and c) countries where relative risk has been calculated with relative risk as a linear scale; grey signifies a country not included.

**Figure 2.**
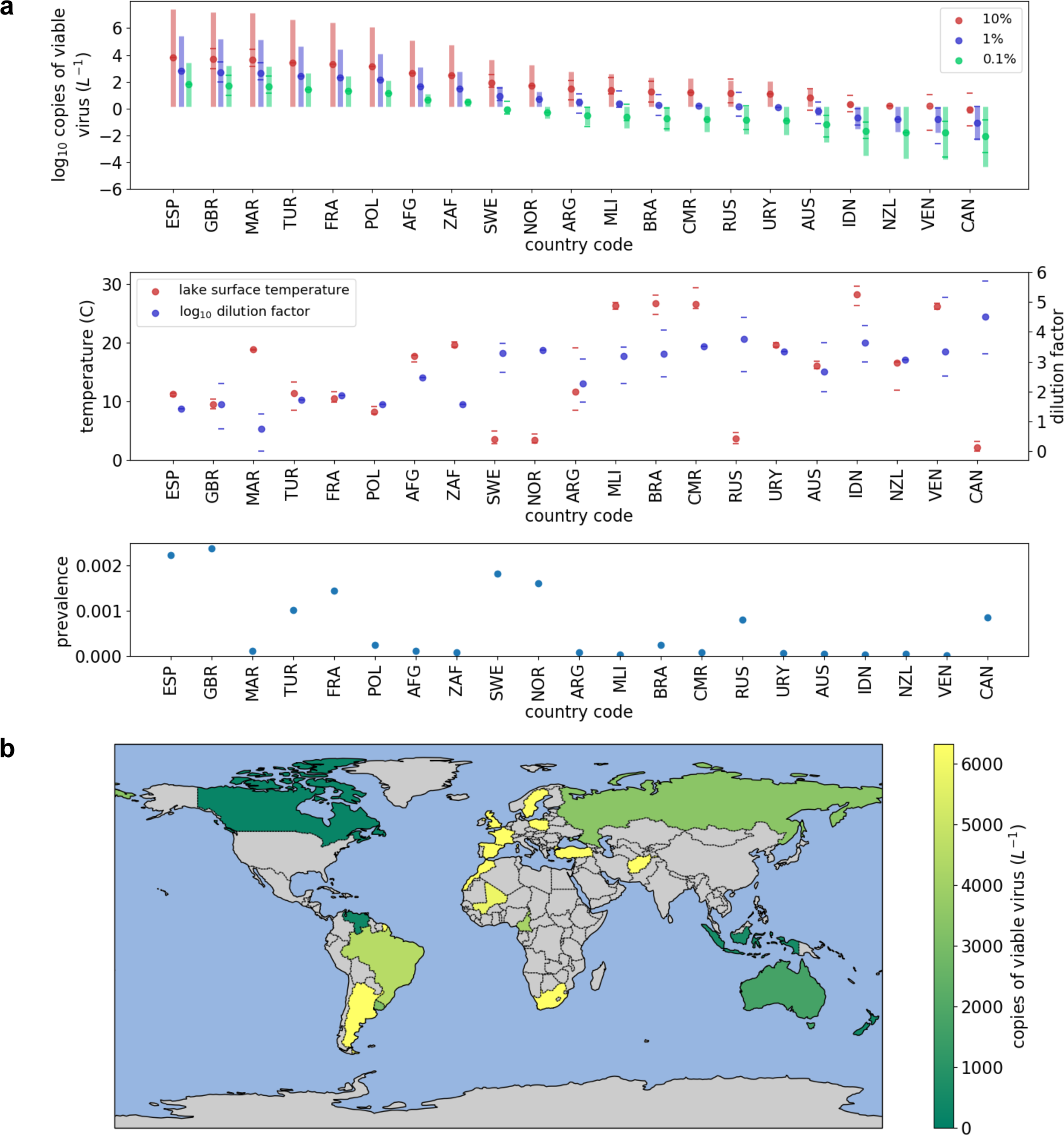
Estimate of absolute viable viral concentration within inland waters on the May 03 2020 for 21 countries assuming a sewage spill has occurred. a) absolute viable viral concentrations in log_10_ copies. Circles are median, horizontal lines are 25^th^ and 75^th^ percentiles due to dilution factors from ^13^; shaded uncertainty bars are ±68% copies L^-1^. Results are shown for three possible ratios of viable virus to viral genome copies (10%, 1% and 0.1%) and b) countries where viable viral loads have been calculated. Grey signifies a country not included; viral concentrations are presented as a linear scale in copies of viable virus.

## Discussion

The detection of SARS-CoV-2 virus in the aquatic environment ^15^ does not necessarily translate into the presence of viable virus. To estimate the number of viable (infectious) virus copies, the proportion of infectious viruses in sewage must be known. The presence of infectious virus in stool samples has been demonstrated ^4^, but there is a lack of quantitative data on this ratio for SARS-CoV-2 in stool. We instead used literature on the number of infectious adenovirus copies in sewage (eg ^16^) and wastewater discharge into rivers ^17^ to select high (10^−1^) medium (10^−2^) and low (10^−3^) estimates for the ratio of infectious virus to genome copies to infectious viruses. We note that adenoviruses are known to be particularly resilient, and therefore likely to represent an upper estimate, but also that our selected range is consistent with the 10^−3^ value used elsewhere for assessing viral risk in water systems (eg ^14^), including one assessment for SARS CoV-2 transmission risk to wastewater workers ^18^.

The temperature dependent survivability means that it is likely that the risk posed by wastewater will increase during winter months as the sewage temperature will be lower enabling longer viral survival, but temperature history and age of the sewage will be needed to fully understand any detected viral loads. SARS-CoV-2 infection to, and spread between, domestic cats has occurred due to similarities between human and some animal angiotensin converting enzyme 2 (ACE2) gene ^20^. Increased animal foraging can occur downstream from water treatment facilities, relative to upstream, highlighting possible risk of some riparian wildlife infection if feeding occurs after a spill.

### Implications for drinking water

It is possible that SARS-CoV-2 survivability and transport within rivers could impact drinking water supplies in countries where rivers or reservoirs are the primary drinking water sources and where large populations, with little or no sewage treatment, exist close to the water source, such as within refugee camps or shanty towns. Riverine enteric virus transport and catchment accumulation can occur for common viruses (eg ^21^) and under stratified conditions it would be possible for a river plume to enter a reservoir and subsequently exit through the reservoir outlet without mixing with the main body of water. Filtering of water, followed by ultraviolet disinfection or chlorination are the recommended approaches for virus removal from drinking water sources ^22^. Filtering is normally used to remove large particulates. The effective ultraviolet dose for SARS-CoV-2 disinfection appears highly variable and dependent upon the surface to which the virus is attached ^23^. The upper dosage value of 1 Joule (J) cm^-2^ to ensure effective ultraviolet disinfection of SARS-CoV-2 ^23^ is an order of magnitude larger than that typically used (∼40 to 90 mJ cm^-2^) for low volume domestic drinking water treatment. The World Health Organization (WHO) guidelines state that effective chlorination disinfection occurs at residual chlorine concentrations of ≥0.5 mg L^-1 22^, which matches the minimum needed to deactivate SARS-CoV-1 ^24^. However, the actual chlorine dosage used for water treatment can vary, based on country, region, water origin and infrastructure (eg UK guidelines are concentrations of 0.2 to 0.5 mg L^-1^). Collectively this means that if a drinking water source was to become infected with SARS-CoV-2 the standard virus removal and disinfection approaches of ultraviolet exposure and chlorination may not reduce the virus below detectable limits. Reviewing of regional or countrywide drinking water processing approaches is recommended to reduce the potential for SARS-CoV-2 surviving through drinking water processing systems. Boiling of drinking water will result in the virus being deactivated ^22^. Refrigerated food that becomes contaminated (eg through washing or handling) could remain infectious for up to 25 days.

### Implications for the marine environment

The virus remains stable over a range of pH ^3^ and in sterile saline solution at low temperatures ^2^, so it is possible that there is no significant difference in virus temporal survival and infection risk between freshwater and seawater, and SARS-CoV-2 has already been identified within seawater, originating from untreated wastewater ^7^. Bioaccumulation of the SARS-CoV-2 virus by molluscs and other aquatic organisms may occur as bivalves are known to accumulate waterborne viruses including hepatitis, norovirus and avian influenza ^25^.

Multiple cetaceans have very high ACE2 similarity to humans making them susceptible to SARS-CoV-2 infection including harbor porpoises, bottle nosed dolphins, minke whales, orca and pilot whales ^20^. Of particular concern are whales whose throats are exposed to large volumes of water during feeding and who visit coastlines for prey that are known to accumulate around sewage outfalls, such as minke whales feeding on mackerel or orca feeding on chinook salmon. In these instances, the animal could be exposed to a large viral dose, even if the virus is only present within the water in low concentrations. For example, if the riverine viral concentration is low at 1 copie ml^-1^, which is undetectable by PCR (detection limit is >100 copies ml^-1^), then a medium sized whale filtering water during feeding could receive repeated doses of 5.65 million copies every second (see methods for calculation). A seafood market is among the suspected sources for the origin of the SARS-CoV-2 virus, so any viral transmission from land to sea may be a circular process.

## Conclusions

Natural water systems are likely able to act as a transmission pathway for SARS-CoV-2 which poses a threat to human infection. The analysis suggests that public interactions with rivers and coastal waters following wastewater spills should be minimized to reduce the risk of infection. New volume integrating viral detection methods are needed to ensure the safety of water systems. While the primary risk associated with the current COVID-19 outbreak appears to be human-to-human transmission of SARS-CoV-2, this work supports the plausibility that novel coronaviruses may also spill over to new wildlife hosts through infected faecal matter accidentally entering the natural aquatic environment; this potential virus reservoir could enable future resurgence in the human population.

## Data Availability

All of the input data are open source and freely available on line.

## Supplementary

Data files are provided for the viable viral counts (viral_counts.xlsx) and relative risk data (relative_risk.xlsx).

## Acknowledgements

JDS, KZ and RD were partially supported by the European Union Marie Curie Innovative Training Network AquaSense (grant H2020-MSCA-ITN-2018-813680). RD was partially supported through Engineering and Physical Science Research Council (EPSRC) Engineering Fellowship for Growth (grant EP/R029644/1). JDS and TH were partially supported by the European Space Agency project OceanSODA (grant 4000125955/18/I-BG).

## Author contributions

JDS, KZ, RD and TH developed the initial ideas. JDS, KZ, TH, MM, KU and LM developed the methods. IA provided computing facilities. All authors contributed to the writing of the manuscript.

## Methods

### Risk from wastewater spillage between countries

The relative risk of SARS CoV-2 from waste water systems is calculated by using a modified version of equations 1 and 2 from ^13^, given as

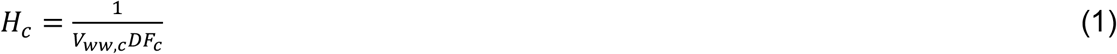

where V_ww,c_ is the per capital daily volume of domestic water usage for country *c*, and DF_c_ is the dilution factor downloaded from ^13^ supplemental table 1 and supplemental table 2, respectively. Normalising H_c_ across all country median DF values provides the between country relative risk of water borne infection due to the viral load in a river following a sewage effluent spill (shown in figures 1b and 1c).

The number of infectious virus copies in the water system as a result of a waste water spill or leak is calculated by multiplying H_c_ by the number of infectious viruses in faeces generated by the infectious proportion of a county’s population, C_inf,c_. This is calculated using

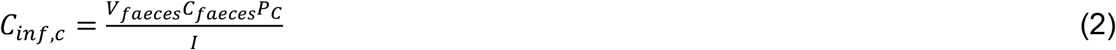

where V_faeces_ is the volume of faeces generated (litres, L, per capita per day), C_faeces_ is the number of viral RNA copies in faecal matter (L^-1^), P_c_ is the proportion of the population of country *c* that have active infections, and *I* is the ratio of viral RNA copies to viable (infectious) virus.

We note that measured wastewater viral counts in Paris on the 9^th^ April were 3.1 × 10^6^ genome copies L^-1^ with 82,000 active cases ^19^, whereas using our (albeit country specific) method gives the estimate of 1.3 × 10^6^ genome copies L^-1^, which is within the correct order of magnitude (this calculation used the same number of active cases).

To calculate C_faeces_ we assumed a log-normal distribution and calculated the expected value using the mean and standard deviation from ^12^ using the standard equation:

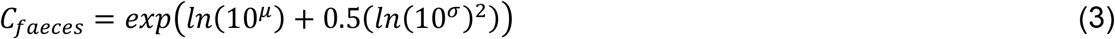

Where µ is the sample mean and σ is the sample standard deviation of the log normal distribution. ^12^ state that µ of the distribution is 5.22 log_10_ copies ml^-1^ and σ = 1.86 log_10_ copies ml^-1^ which results in an expected C_faeces_ concentration within the sewage effluent of 1595.9 million copies ml^-1^. V_faeces_ is the mean daily volume of faeces generated per person (0.149 kg, from table 3 of ^26^ and assuming faeces has a density approximately equal to water ^27^. Note we used the ‘rich country’ value from ^26^ because the RT-PCR data ^12^ that we use to estimate C_faeces_ was measured from samples collected in Germany. The prevalence data, P_c_, were calculated by subtracting the number of recovered and number of fatalities from the number of confirmed cases from the Worldometer website.

PCA does not distinguish between infectious virus and damaged/destroyed non-infectious virus. Therefore, to estimate the number of viable (infectious) virus copies, we used literature on the ratio of infectious adenovirus copies to genome copies in raw sewage (eg Rodríguez et al., 2013) wastewater discharged into rivers ^17^. These estimates varied over four orders of magnitude, and as such we selected high (10^−1^), medium (10^−2^) and low (10^−3^) estimates (which equate to 10%, 1% and 0.1% proportion of viable versus within the total viral genome counts).

The expected number of copies of infectious virus resulting from a sewage spill into a river, lake or coastal region for a given country can therefore be calculated as

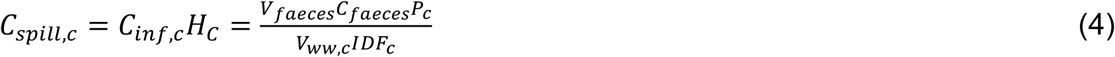

C_spill,c_ was estimated for May 3 2020 21 countries that contain large inland water bodies and were known to rely upon reservoirs for drinking water ^28^. Long-term statistical mean water temperature, needed to calculate virus survivability, was calculated from a climate quality global lake temperature dataset (see below). Temperature values for each country were the countrywide mean lake temperature within a rectangular box matching a simplified country outline. The dilution factors reported in ^13^ can vary by several orders of magnitude and were deemed to provide the major source of uncertainty in the calculation. Therefore, the C_spill,c_ viral loadings given by the 25^th^ percentile dilution, median dilution and 75^th^ percentile dilution values are all presented. With high, medium and low estimates for I, this results in nine estimates of C_spill_ for each country.

The long-term statistical mean global lake water temperature climatology was constructed using the 0.05° × 0.05° daily resolution GloboLakes v4 data set ^29^ which covers 1996 to 2016. Mean temperature was calculated for each calendar month across all years producing 12 monthly mean datasets with a 0.05° × 0.05° gridded resolution. Uncertainty terms were propagated by assuming random errors were independent and normally distributed, and using standard error propagation methods. The resulting uncertainty term combines the original uncertainty in measurement and optimum interpolation with the spatial/temporal uncertainty of the resampled monthly average, for each grid cell.

The concentration of SARS-CoV-2 virus needed for infection is not known. ^30^ provides 10^3^ copies for influenza. The Infectious dose for SARS-CoV-2 is likely significantly lower because ^31^ ranks influenza as “very high infective dose” and SARS-CoV-2 as “low”. We therefore use a value of 100 copies as a concentration that could result in infection.

A combined uncertainty budget for equation 4 was calculated using standard uncertainty propagating methods and estimates of the uncertainties of each input dataset. Uncertainty components (and their values) were domestic water usage (±10%), population size (±1%), number of active cases (±20%), mass of faeces generated per capita per day (0.095 kg, see table 3 of ^26^, mean number of viral genome copies in faeces (3.54×10^12^) and density of faeces was not included in the uncertainty analysis. This resulted in a combined uncertainty budget of ±68% copies ml^-1^. It is important to note that this value does not include uncertainty in the dilution factors or the ratio of viral genome copies to infectious virus. Instead, the C_inf_ calculation was repeated for high, medium and low values of these parameters.

### Temperature dependent survival

As reported in ^3^, the virus concentration in water follows an exponential decay, with its half-life decreasing with decreasing temperature and the pH control of half life is very small over the pH range of 3-10 (which encompasses the range found in natural freshwater and marine systems). Based on the in vitro data presented in ^3^, the following empirical model was derived to describe virus concentration reduction factor due to the temperature-dependent die-off:

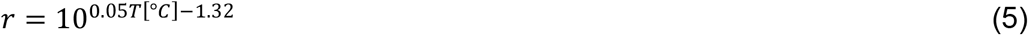

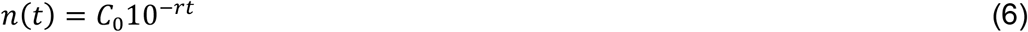

Where C_0_ is initial virus concentration (copies ml^-1^), *n(t)* is virus concentration after time *t* (days) and *r* is 24 hour survival factor due to temperature *T* driven die off. This model fit to the in vitro data gives a root mean square difference (RMSD) of ±1% for water at 4°C which increases to ±7.5% at 22°C. When considering temperature controlled survival in the waste water system, C_ww,c_ becomes the value used for the initial viral load *C*_*0*_ following a sewage effluent spill. As noted in ^5,12^, the viral load follows a heavy-tailed distribution with the majority of patients shedding around 10^5^ copies ml^-1^) but some having viral loads as high as 10^12^ copies ml^-1^. This results in the super-spreader problem where a tiny proportion of the infected population can become responsible for contributing a majority of viral load in the wastewater. For a large infected population, this approach allows robust statistical modeling of viral load. However, in case of smaller communities with low number of infections, the actual viral load could be severely underestimated if a super-spreader is present within the population.

### Whale filtering calculation

The example volume flow rate through the mouth of a medium sized Bowhead whale whilst feeding was provided by ^32^) A flow rate of 5.65 m^3^ s^−1^ is given for a 15 m whale (mouth pressure of –1768 Pa at a 4 km h^−1^ foraging speed, assuming an oral opening of 5.09 m^2^ with an opening radius = 1.27 m). Assuming a low viral concentration of 1 copies per ml^-1^, which equates to 1000 copies l^-1^. 5.65 m^3^ s^−1^ equates to 5650 L s^-1^. The dosage per second as the whale swims during feeding is given by 1000 (copies L^-1^) × 5650 (L s^-1^) = 5.65 million copies s^-1^.

